# The impact of physical distancing measures against COVID-19 transmission on contacts and mixing patterns in the Netherlands: repeated cross-sectional surveys in 2016/2017, April 2020 and June 2020

**DOI:** 10.1101/2020.05.18.20101501

**Authors:** Jantien A. Backer, Liesbeth Mollema, R.A. Eric Vos, Don Klinkenberg, Fiona R.M. van der Klis, Hester E. de Melker, Susan van den Hof, Jacco Wallinga

## Abstract

**Background:** During the current COVID-19 pandemic many countries have taken drastic measures to reduce the transmission of SARS-CoV-2. These often include decreasing the number of contacts by physical distancing.

**Aim:** To measure the actual reduction of contacts when physical distancing measures are implemented.

**Methods:** In the Netherlands, a cross-sectional survey was carried out in 2016/2017 in which participants reported the number and age of their contacts during the previous day. The survey was repeated among a subsample of the participants in April 2020 after physical distancing measures had been implemented, and in an extended sample in June 2020 after some of these measures were relaxed.

**Results:** The average number of community contacts per day was reduced from on average 14.9 (interquartile range: 4-20) in the 2016/2017 survey to 3.5 (0-4) after physical distancing measures were implemented, and rebounded to 8.8 (1-10) after some of these measures were relaxed. All age groups restricted their number of community contacts to at most 5 contacts on average after physical distancing measures were implemented. After relaxation, children reverted to baseline levels while elderly had a number of community contacts that was less than half their baseline levels.

**Conclusion:** The physical distancing measures have greatly reduced contact numbers, which has likely been beneficial in curbing the first wave of the COVID-19 epidemic in the Netherlands. Different age groups reacted differently upon relaxation of these measures. These findings offer guidance for the deployment of age-targeted measures in the future course of the pandemic.

## Introduction

Since the beginning of 2020, the novel coronavirus SARS-CoV-2 that causes COVID-19 disease has rapidly spread around the world. Most patients experience mild symptoms, but in particular elderly and persons with comorbidities may develop severe acute respiratory disease [1]. Hospitals have been confronted with an alarmingly growing number of patients, often exceeding the intensive care (IC) capacity. Many countries have implemented control measures that include a combination of increased hygiene, travel restrictions, case finding, contact tracing and physical distancing measures. The specific physical distancing measures differ between countries and between regions; their overall aim is to reduce the number of contacts in the population thus preventing the transmission of infection. The impact of these measures on the reduction of contacts in the population, and the reduction of contacts made by different age groups within that population, are poorly quantified.

Different approaches exist to measure behavioural changes. Mobile telephone data provided by telecom companies are used to measure the change in mobility patterns [2, 3]. Similarly, the location history of smart phones can be tracked with apps [4]. The anonymised and aggregated mobility patterns can suggest changes in contact patterns in the population at large. To obtain direct and detailed information on contacts, cross-sectional studies are conducted in which participants report their age and sex as well as the age and sex of all persons they have contacted during a single day [5, 6]. In the first months of the COVID-19 pandemic, contact surveys have been successfully used to quantify the reduction in the number of contacts associated with physical distancing measures in Shanghai and Wuhan, China, estimated at 88% and 86%, respectively [7], and in the UK among the adult population, at 74% [8]. One of the challenges with the contact survey approach is to obtain a reliable baseline measurement before physical distancing measures were implemented. In the Wuhan study, participants needed to recall the number of contacts during a regular weekday at the end of 2019. In the UK study the baseline was provided by a similar study conducted 13 years ago among a different representative UK study population [5].

We present two large contact surveys conducted in the Netherlands, in April 2020 and in June 2020. The participants were recruited from a large nationwide sample of the Dutch population who had participated in an earlier cross-sectional survey in 2016/2017 [9], supplemented with a new nationwide sample of the Dutch population for the June 2020 survey. The contact questionnaire was nearly identical in all surveys, which allows us to use the first survey as a baseline measurement. By 16 March 2020, the Netherlands had imposed physical distancing measures including closing daycare centers, schools, universities; working from home whenever possible; closing of cafes, pubs, restaurants, theaters, cinemas, and sport clubs; cancelling events with more than 10 persons attending; and maintaining 1.5 m distance from others outside one’s household. A few weeks after these measures were implemented, the first survey was conducted. By 1 June 2020, most of the physical distancing measures were somewhat relaxed, except for working from home, and keeping 1.5 m distance. Primary schools and daycare centers were open at full capacity, and secondary schools, cafes, pubs, restaurants, theaters and cinemas at a reduced capacity. A few weeks after this relaxation, the second survey was conducted.

By comparing the survey results before and after implementing the physical distancing measures, we could determine the impact on the number of contacts made in the community (i.e. outside the household), distinguishing between different age groups, sexes, household sizes, and days of the week. We also assessed how the physical distancing measures have affected the total number of contacts, including contacts with household members, and the age-specific mixing patterns.

## Methods

Between February 2016 and October 2017, a cross-sectional sero-epidemiological study was conducted in a sample of the Dutch population from 0 to 89 years of age [9], henceforth referred to as ‘the baseline survey’. Participants were randomly selected from the Dutch population registry using a two-stage cluster design. Infants under 1 year of age, persons living in areas with low vaccination coverage, and persons with a migration background were oversampled in this survey. The study consisted of an extensive questionnaire, that was filled out by the guardians or parents for participants under 15 years of age. It included questions regarding the participants’ age and sex, the age and sex of their household members, and the total number of unique persons they had contacted outside their household the previous day and which day of the week (i.e., Monday through Sunday) this applied to. Participants reported the age of the contacted persons in 11 age groups (0-4, 5- 9, 10-19, 20-29, 30-39, 40-49, 50-59, 60-69, 70-79, 80-89, 90+). In the questionnaire, examples of a contact are given as talking to someone (face-to-face), touching someone, kissing someone or doing sports with someone.

Of the 8,179 participants that had filled out the questionnaire in 2016/2017, 6102 were invited on 26 March 2020 to participate in the follow-up study, referred to as ‘the April 2020 survey’. In total, 3,168 questionnaires were returned. Of these participants 2,754 again participated in the final survey, referred to as ‘the June 2020 survey’. They were supplemented with 4,496 new participants from a large population-based sample of 27,053 randomly drawn Dutch citizens across all municipalities of the country. These bring the total number of participants for the June 2020 survey to 7,250.

In order to obtain a representative study population, we omitted the participants from the low vaccination coverage areas that were oversampled, and we omitted the oversampled infants under 1 year of age in the baseline survey, such that the fraction of infants in the study population reflected the fraction of infants in the 2017 Dutch population. We excluded participants that did not fill out certain questions: participants that did not report the household composition and participants that did not report any contacts while also omitting the contact day. Finally, participants reporting more than 100 contacts per day were excluded from the analysis, as it was deemed unrealistic to have more face-to-face conversations during one day.

The questionnaire in the 2020 surveys was identical to the baseline survey questionnaire, apart from two questions. A question asking whether participants had had any contacts outside their household was added (before asking how many), and in the June 2020 survey a question was added how many contacts in each age group occurred within 1.5 m or outside 1.5 m distance.

We analysed the contact surveys by comparing the number of contacts in the community per participant stratified by several characteristics: age, sex, household size, and day of the week. We combined the two oldest age groups and used 10 age groups (up to 80+) for the analysis. For the June 2020 survey, we studied the fraction of close contacts (within 1.5 m) by age group. We restricted this part of the analysis to community contacts, i.e. contacts made with non-household members, because contacts with household members were not reported.

For the following part of the analysis we used the household composition as a proxy for household contacts. By adding these to the reported community contacts, we obtained the total number of contacts in the population. We estimated age-stratified contact matrices that contain the number of contacts made between and within age groups, using an approach that accounts for reciprocity of contacts between different age groups [10], and using age-specific population size data for the Netherlands on 1 January 2017 and on 1 January 2019 [11]. To check the effect of enforcing reciprocity between contacts, we compared the estimated and observed mean number of contacts per participant. We characterised the mixing pattern of the age-specific contacts by the disassortativeness index [12], which indicates assortative mixing for values of 0 and random mixing for values of 1. We characterised the ‘effective number’ of age-specific contacts by the largest eigenvalue of the contact matrix [13]. All analyses were done using R version 3.6.0 [14].

## Results

### Characteristics of the study population

In total 5066 participants of the baseline survey, 2069 participants of the April 2020 survey, and 6300 participants of the June 2020 survey were included for analysis. The composition of the survey population by age and sex should reflect the Dutch population (Fig. 1), but the older age groups were overrepresented in the 2020 surveys. The mean age was 37 years (range 0 – 88) in the baseline survey, 42 years (range 3 – 90) in the April 2020 survey, and 46 (range 1 – 90) in the June 2020 survey, whereas the mean age of the Dutch population is 41 years. In all surveys, each age group consisted of more than 100 participants, except for the 80+ age group in the baseline survey, and the 0-4 and 80+ age groups in the April 2020 survey (Tab. 1). As expected from the household size distribution in the Netherlands [15], participants lived mostly in 2-person households, followed by 4-person households. Although the April 2020 survey population contained relatively few single-person households, the reported average household size was similar across all surveys (2.8 for the baseline survey, 3.0 for the April 2020 survey, 2.9 for the June 2020 survey), and in line with the expected average household size of 2.8. There were at least 79 participants per contact day of the week for each survey.

**Table 1.** Participant characteristics for the baseline survey in 2016/2017, the April 2020 survey, and the June 2020 survey in the Netherlands.

|  | **baseline** | **April 2020** | **June 2020** |
| --- | --- | --- | --- |
|  | n (%) | n (%) | n (%) |
| Total | 5,066 (100) | 2,069 (100) | 6,300 (100) |
| Participant age group |  |  |  |
| 0-4 | 377 (7.4) | 39 (1.9) | 118 (1.9) |
| 5-9 | 321 (6.3) | 110 (5.3) | 256 (4.1) |
| 10-19 | 597 (11.8) | 194 (9.4) | 575 (9.1) |
| 20-29 | 781 (15.4) | 275 (13.3) | 662 (10.5) |
| 30-39 | 689 (13.6) | 331 (16.0) | 736 (11.7) |
| 40-49 | 613 (12.1) | 312 (15.1) | 859 (13.6) |
| 50-59 | 563 (11.1) | 318 (15.4) | 1028 (16.3) |
| 60-69 | 647 (12.8) | 279 (13.5) | 1163 (18.5) |
| 70-79 | 397 (7.8) | 179 (8.7) | 770 (12.2) |
| 80+ | 81 (1.6) | 32 (1.5) | 133 (2.1) |
| Participant sex |  |  |  |
| Female | 2,803 (55.3) | 1,156 (55.9) | 3,490 (55.4) |
| Male | 2,263 (44.7) | 913 (44.1) | 2,810 (44.6) |
| Household size* |  |  |  |
| 1 | 882 (17.4) | 168 (8.1) | 700 (11.1) |
| 2 | 1,844 (36.4) | 727 (35.1) | 2,499 (39.7) |
| 3 | 667 (13.2) | 359 (17.4) | 902 (14.3) |
| 4 | 1,068 (21.1) | 562 (27.2) | 1,496 (23.7) |
| 5 | 451 (8.9) | 197 (9.5) | 543 (8.6) |
| 6+ | 154 (3.0) | 56 (2.7) | 160 (2.5) |
| Contact day |  |  |  |
| Monday | 918 (18.1) | 521 (25.2) | 1,113 (17.7) |
| Tuesday | 831 (16.4) | 546 (26.4) | 1,047 (16.6) |
| Wednesday | 517 (10.2) | 368 (17.8) | 1,004 (15.9) |
| Thursday | 264 (5.2) | 205 (9.9) | 672 (10.7) |
| Friday | 428 (8.4) | 102 (4.9) | 367 (5.8) |
| Saturday | 683 (13.5) | 79 (3.8) | 746 (11.8) |
| Sunday | 980 (19.3) | 246 (11.9) | 1,348 (21.4) |
| (Missing) | 445 (8.8) | 2 (0.1) | 3 (0.0) |

\* Households in the Netherlands consist of 1 (38%), 2 (33%), 3 (12%), 4 (12%) and 5+ (5%) persons [15], leading to an expected distribution per participant to live in a 1 (17%), 2 (31%), 3 (17%), 4 (23%) or 5+ (12%) person household.

**Figure 1.**
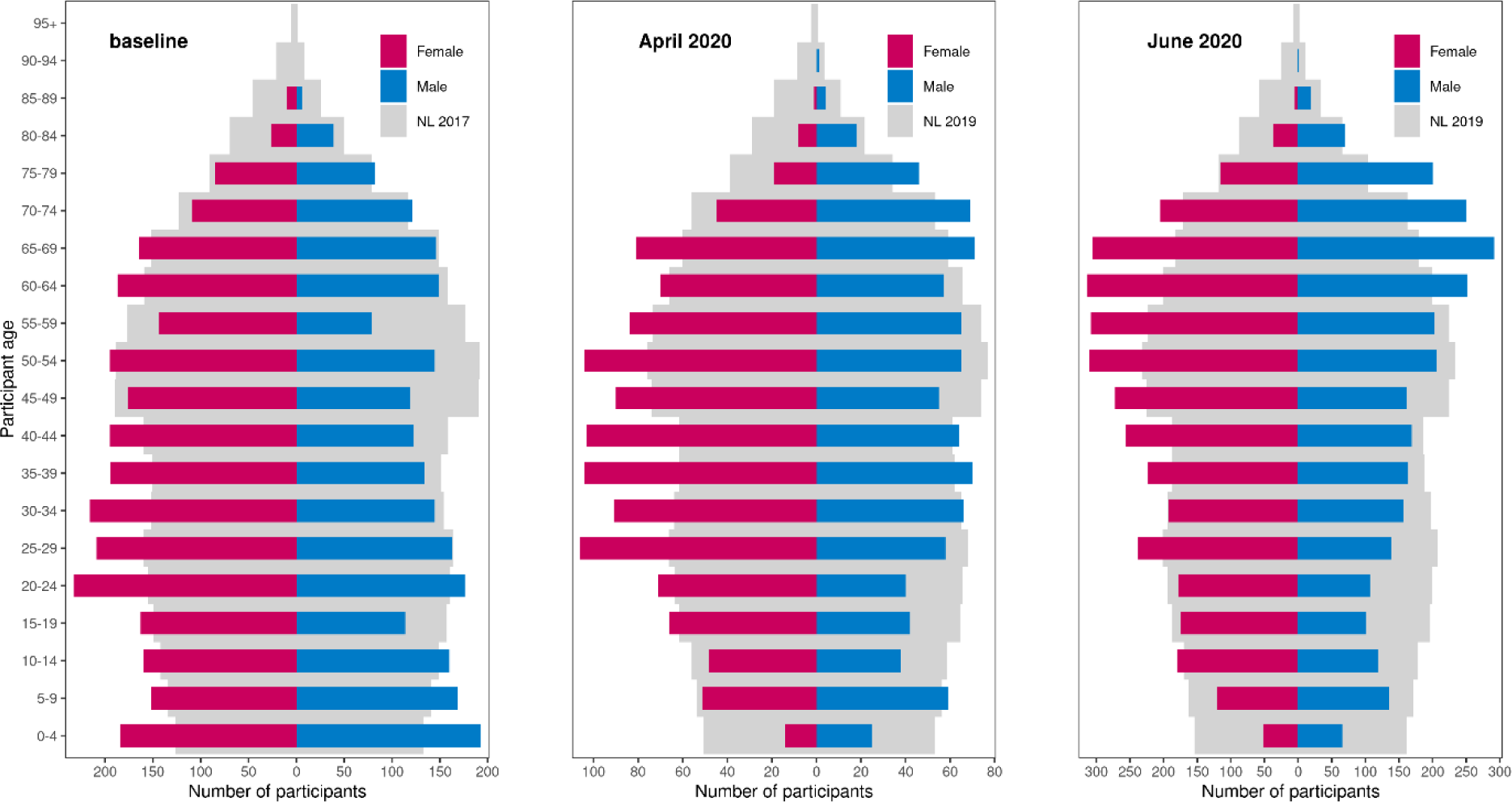
Composition according to age and sex of 5,066 participants in the baseline survey in 2016/2017, 2,069 participants in the April 2020 survey, and 6,300 participants in the June 2020 survey in the Netherlands; compared to the Dutch population in 2017, 2019 and 2019, respectively.

### Reduction in the mean number of community contacts

The percentage of participants who did not report any community contacts on a single day increased from 5% in the baseline survey to 42% in the April 2020 survey, and decreased again to 22% in the June 2020 survey. The average number of community contacts a participant reported per day, decreased from 14.9 (4 – 20, interquartile range or IQR) in the baseline survey to 3.5 (0 – 4) in the April 2020 survey, and rebounded to 8.8 (1 – 10) in the June 2020 survey (Tab. 2). Compared to the baseline survey, the number of community contacts reduced by 76% and 41% in the April 2020 and June 2020 surveys respectively.

**Table 2.** Number of community contacts per participant in the baseline survey in 2016/2017, the April 2020 survey, and the June 2020 survey in the Netherlands (mean and interquartile range). The relative change in mean number of community contacts compared to the baseline survey is shown in shades of red (for reduction) and blue (for increase).

|  | baseline |  | April 2020 |  | June 2020 |  |
| --- | --- | --- | --- | --- | --- | --- |
|  | mean | (IQR) | mean | (IQR) | mean | (IQR) |
| Total | 14.9 | (4 - 20) | 3.5 | (0 - 4) | 8.8 | (1 - 10) |
| Participant age |  |  |  |  |  |  |
| 0-4 | 13.8 | (3 - 19) | 3.7 | (0 - 4) | 18.0 | (3 - 26) |
| 5-9 | 22.1 | (6 - 33) | 2.1 | (0 - 3) | 27.1 | (6 - 39) |
| 10-19 | 22.5 | (7 - 33) | 2.9 | (0 - 4) | 14.3 | (1 - 24) |
| 20-29 | 16.6 | (5 - 22) | 3.6 | (0 - 5) | 9.4 | (1 - 11) |
| 30-39 | 14.3 | (5 - 18) | 4.2 | (0 - 6) | 9.7 | (2 - 12) |
| 40-49 | 15.3 | (4 - 20) | 4.8 | (0 - 5) | 9.3 | (2 - 11) |
| 50-59 | 13.0 | (4 - 16) | 4.7 | (0 - 6) | 7.1 | (1 - 9) |
| 60-69 | 9.7 | (3 - 11) | 2.2 | (0 - 3) | 5.9 | (0 - 6) |
| 70-79 | 7.8 | (2 - 9) | 1.8 | (0 - 2) | 2.7 | (0 - 4) |
| 80+ | 7.9 | (2 - 8) | 0.7 | (0 - 1) | 3.2 | (0 - 4) |
| Participant sex |  |  |  |  |  |  |
| Female | 14.3 | (4 - 19) | 3.4 | (0 - 4) | 8.9 | (1 - 10) |
| Male | 15.6 | (4 - 22) | 3.7 | (0 - 4) | 8.7 | (1 - 10) |
| Household size |  |  |  |  |  |  |
| 1 | 12.7 | (3 - 16) | 3.1 | (0 - 3) | 5.2 | (0 - 6) |
| 2 | 13.2 | (3 - 17) | 3.0 | (0 - 3) | 5.8 | (0 - 7) |
| 3 | 13.9 | (4 - 19) | 4.5 | (0 - 5) | 10.1 | (1 - 12) |
| 4 | 17.4 | (5 - 25) | 3.7 | (0 - 4) | 12.1 | (2 - 15) |
| 5 | 19.2 | (6 - 29) | 3.7 | (0 - 5) | 13.5 | (2 - 18) |
| 6+ | 20.5 | (6 - 30) | 3.1 | (0 - 4) | 16.5 | (2 - 24) |
| Contact day |  |  |  |  |  |  |
| Monday | 16.0 | (4 - 22) | 3.5 | (0 - 4) | 10.4 | (1 - 12) |
| Tuesday | 17.1 | (5 - 25) | 3.5 | (0 - 4) | 10.0 | (1 - 12) |
| Wednesday | 15.8 | (5 - 21) | 3.9 | (0 - 4) | 9.3 | (1 - 11) |
| Thursday | 17.0 | (4 - 24) | 4.3 | (0 - 5) | 9.4 | (1 - 11) |
| Friday | 16.3 | (4 - 22) | 3.6 | (0 - 4) | 13.6 | (2 - 16) |
| Saturday | 12.9 | (4 - 17) | 4.2 | (0 - 4) | 8.1 | (2 - 9) |
| Sunday | 10.1 | (3 - 12) | 2.4 | (0 - 3) | 4.8 | (0 - 5) |
| (Missing) | 18.1 | (5 - 25) | 3.0 | (3 - 3) | 21.0 | (2 - 31) |

In the baseline survey, participants aged 10 to 19 years had the highest number of community contacts, and this number gradually declined with increasing age. In contrast, the number of community contacts was more similar across the different age categories in the April 2020 survey with middle-aged participants (30-59 age groups) reporting the highest numbers of contacts (Fig. 2). The reduction in the number of contacts was with 90% greatest for participants aged 5 to 9 years, and with 64% lowest for participants aged 50 to 59 (Tab. 2). After measures were relaxed, contact numbers increased among all age groups. Compared to the baseline survey, the two oldest age groups still reduced their contacts by 65% and 59%, but the two youngest age groups increased the number of contacts by 30% and 23%.

**Figure 2.**
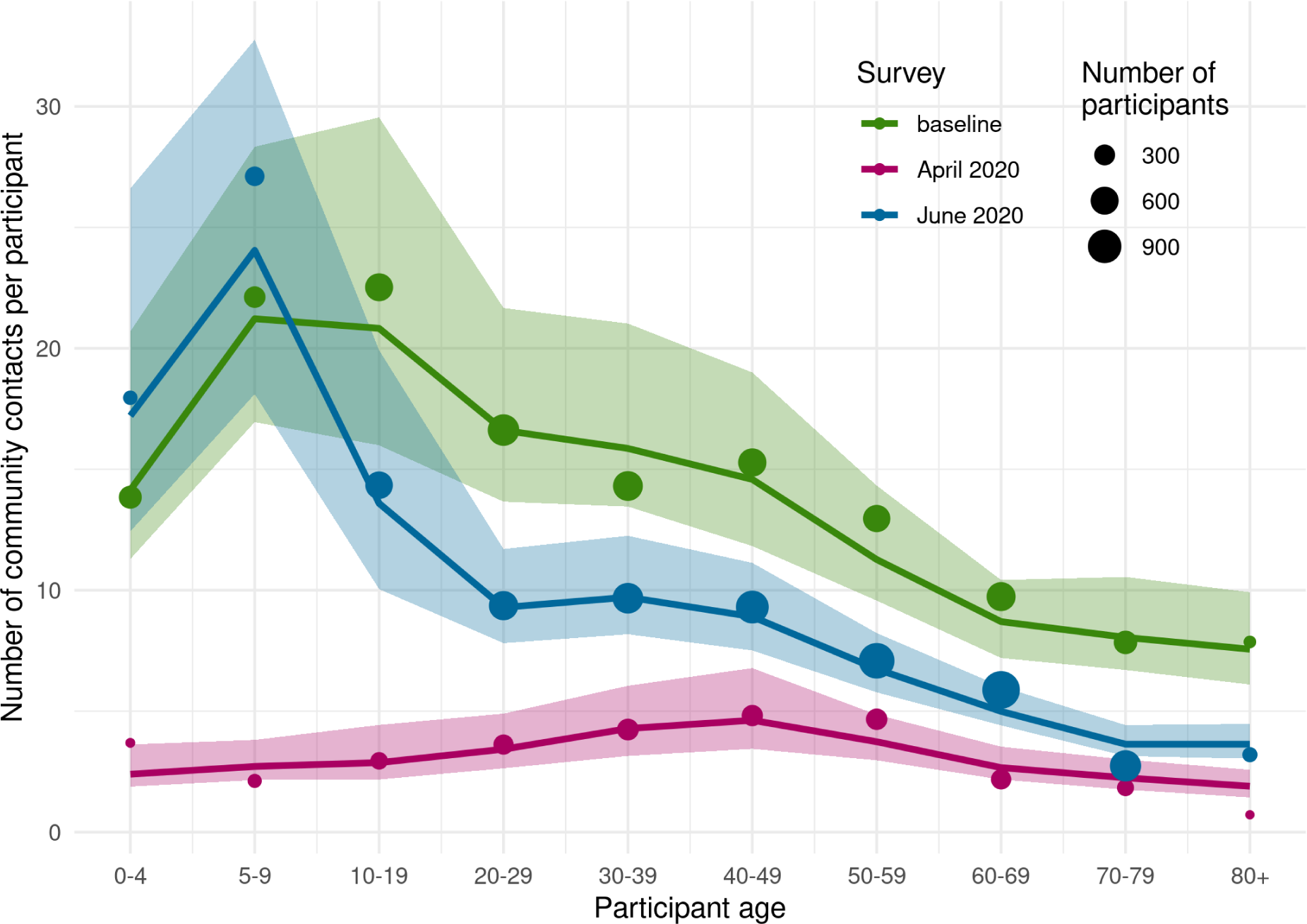
Number of community contacts per participant per age group in the baseline survey in 2016/2017 (green), the April 2020 survey (red), and the June 2020 survey (blue) in the Netherlands; shown are the estimated mean numbers of contacts after accounting for reciprocity in contacts (line) with the corresponding 95% credible interval (shaded area), and the observed means (size of points scaled to number of participants).

The reduction in the mean number of contacts was similar for male and female participants. The number of community contacts increased with household size in the baseline survey, whereas this number was similar for the different household sizes in the April 2020 survey. In the June 2020 survey, the number of community contacts increased for all household sizes, albeit to a lesser extent in 1- and 2-person households. This is likely caused by elderly persons who primarily live in smaller households (see S1.1). In the baseline survey, most contacts were made during weekdays, and fewer in the weekends. This distinction largely disappeared during the physical distancing measures, and reappeared after measures were relaxed.

In the last contact survey, a question was included whether contacts occurred within or outside 1.5 m distance. On average, 53% of the community contacts per participant were reported within 1.5 m, ranging from 44% for the 70-79 age group to 77% for the 0-4 age group (Fig. 3). These close contacts outside the household occur mainly between children themselves, and between 0-4 year-olds and 50-79 year-olds (see S1.2).

**Figure 3.**
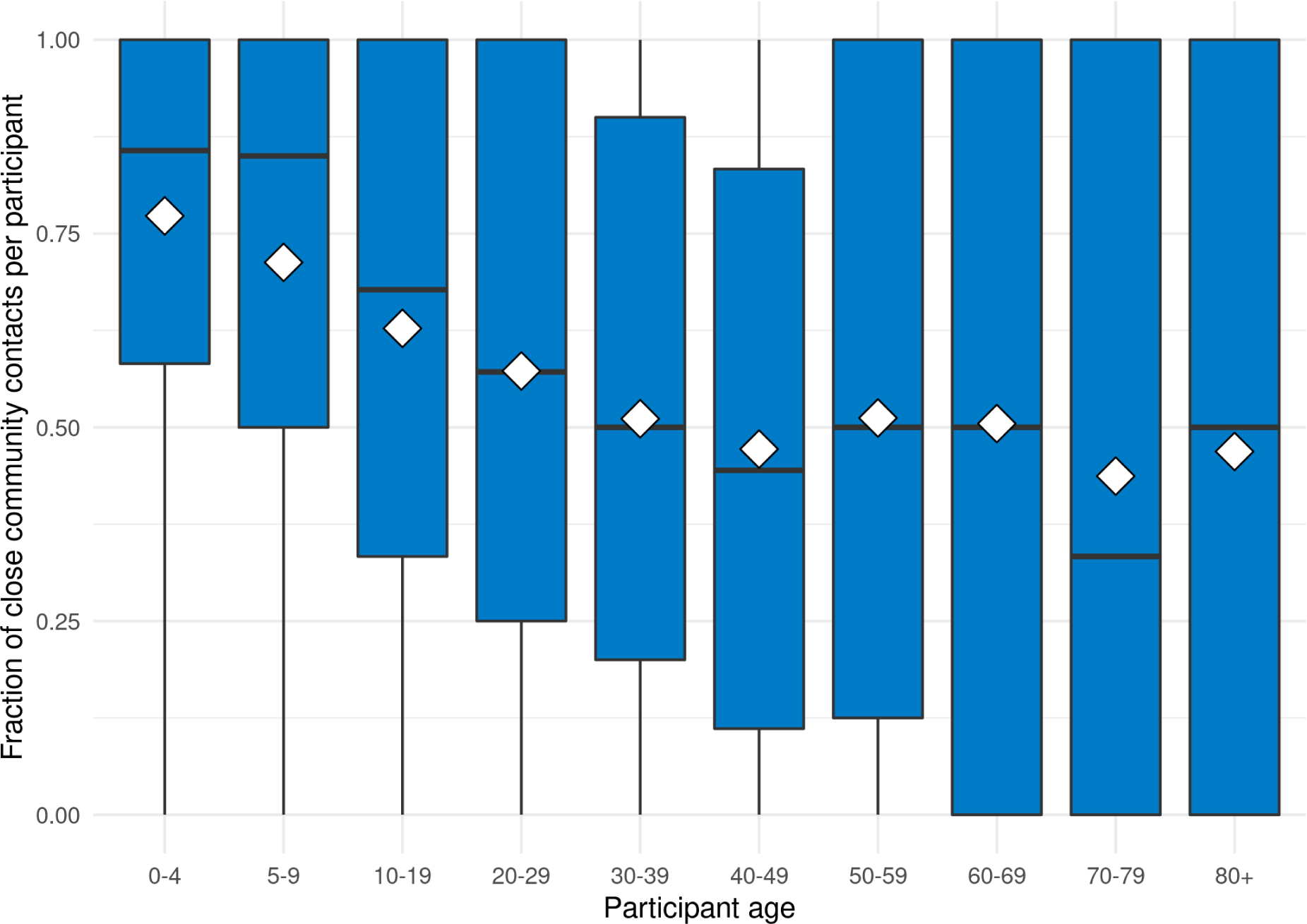
Fraction of close community contacts per participant for different age groups, as observed in the June 2020 survey, with mean (white diamond), median (black line), and interquartile range (blue area). Close contacts were defined as occurring within 1.5 m.

### Comparison of mixing patterns

In estimating the contact rates between age groups, reciprocity between contacts was explicitly taken into account. The observed (points) and estimated (line) mean numbers of community contacts per participant are in agreement (Fig. 2), showing consistency in reporting contacts between the age groups.

The contact matrices for all contact surveys and contact types are shown in Fig. 4. The contact matrices for household members illustrate that participants generally live with persons in their own age group and with their children or parents (i.e. 30 years younger or older); this is apparent in all surveys as the household composition remains fairly constant over time. The matrices for contacts in the community indicate the younger age groups have fewer contacts with all other age groups in the April 2020 survey compared with the baseline survey. Most community contacts are made among the working age classes and between elderly (80+) and middle-aged adults who might be health care workers or informal care givers. In the June 2020 survey, the community contact pattern is restored to the baseline pattern, but the absolute values of the contact numbers are smaller.

**Figure 4.**
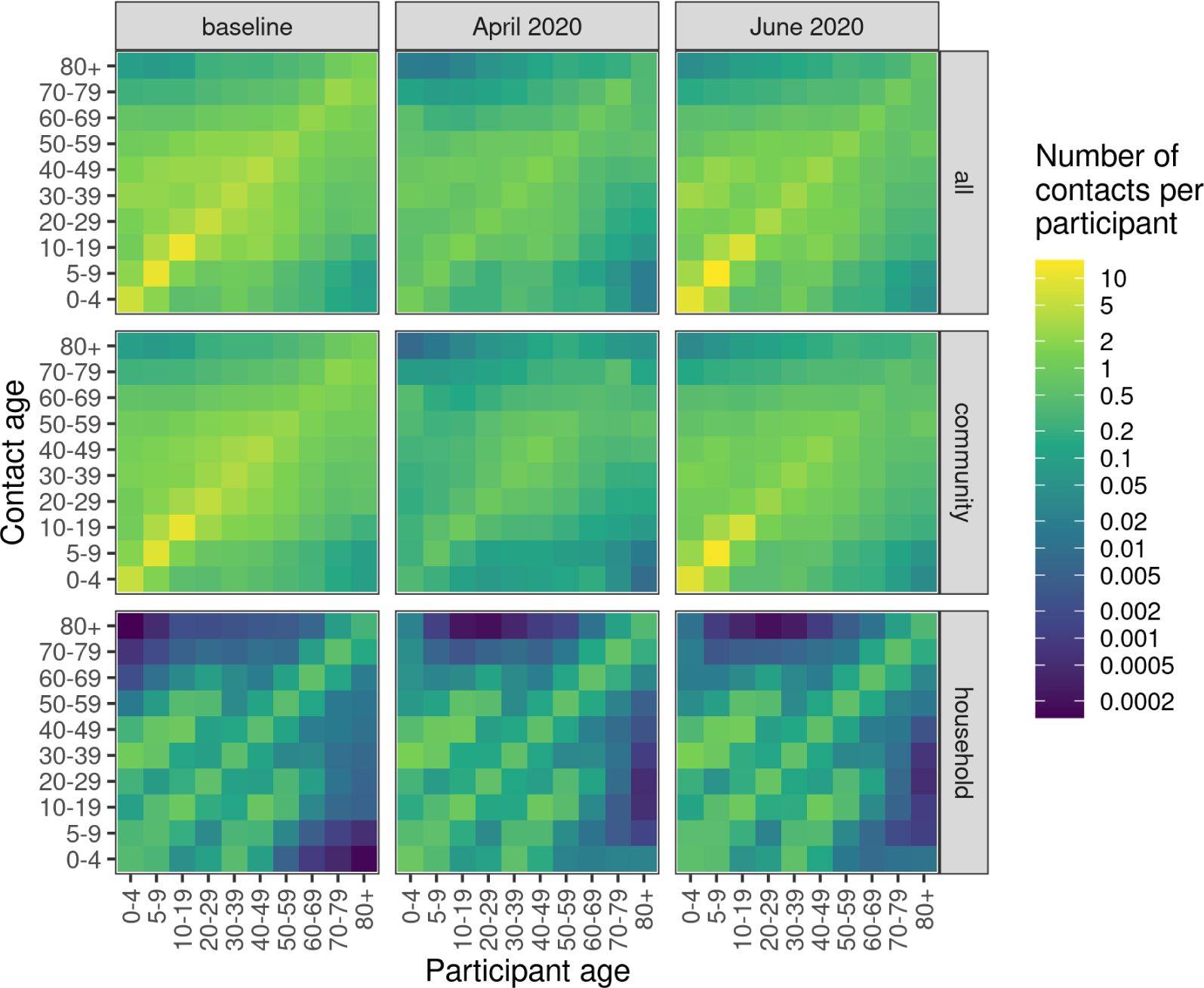
Estimated contact matrices for the baseline survey in 2016/2017, the April 2020 survey, and the June 2020 survey in the Netherlands, for all contacts, contacts in the community and contacts with household members.

Taken together, the household and community contact patterns reveal that age-specific mixing did not change much between the baseline and the April 2020 surveys (disassortativeness index is 0.50 for the baseline survey, 0.52 for the April 2020 survey). For the June 2020 survey however, the disassortativeness index is 0.66, indicating that mixing shifted slightly from within same age groups to between different age groups. The effect would be that a virus would spread more easily in the population. This is reflected in the effective number of contacts per person (i.e. the largest eigenvalue of the contact matrix), which decreased from 18 (15 – 24, 95% credible interval) in the baseline survey to 5.6 (4.7 – 6.5) in the April 2020 survey, an average reduction of 69% (58% – 77%), and reverted to 18 (14 – 27) the June 2020 survey.

## Discussion

In the Netherlands, physical distancing measures came into effect on 16 March 2020. Three weeks after implementation, the numbers of occupied hospital and IC beds reached a peak, after which they gradually declined, reaching a minimum by June 2020 when most measures had been relaxed to some degree. Through comparison with the baseline survey, the numbers of contacts in the community were reduced by 76% by the physical distancing measures, and by 41% after relaxing the measures.

The sampling scheme for inviting participants from the Dutch population was designed to obtain a representative study population. To assess the representativeness of the survey participants, a few potential limitations need to be addressed. First, not all of those who were invited to the survey, participated, causing a potential for selection bias. Second, there are differences in participant characteristics between the surveys, for instance participant age, sex, household size and contact day. We determined the weighted average number of contacts to account for these differences (see S1.3), and found that only the participant age altered the unweighted averages by about 10%. This does not affect the results as they are all stratified by age. Third, the 2020 surveys were carried out in time windows of one month (April and June), whereas the baseline survey was conducted over a period of almost two years. Because contact patterns change little throughout the course of a year (see S1.4 and [16]), we do not expect this to substantially affect the estimated reduction. Finally, the surveys consisted of different but overlapping study populations. To check whether this had any effects on the results, we repeated the analysis on 1,739 participants that were common to all surveys. This longitudinal study showed that – although baseline levels were a bit higher – trends and reductions were similar to the main analysis (see S1.5).

During strict physical distancing, the number of community contacts was drastically reduced in all age groups. When these measures were relaxed, elderly persons largely kept their contacts at a reduced level, while children under 10 years of age had a number of contacts similar to before the measures were implemented. Moreover, the majority of these contacts occurred occurred within 1.5 m, whereas in the rest of the population around half of the contacts were made in close range. On the population level, children under 10 years of age accounted for 23% of all contacts (compared to 13% in the baseline survey) but only for less than 1% of the reported infections [17]. This confirms the minor role that children play in the epidemic, as is also found in age-specific seroprevalence studies [18-20].

The reduction in the number of contacts associated with physical distancing measures in the Netherlands is smaller than the reductions of 88% and 86% observed in Shanghai and Wuhan, China [7], most probably because the measures in the Netherlands were less stringent than those in both Chinese cities. In the UK a 74% reduction in the number of all contacts among adults (18+) has been reported [8]. To compare this with our results, we calculated the total number of contacts including community and household contacts for the participants of 18 years and older in our surveys, and found a reduction of 72%. At the time of the studies the control measures in the UK and the Netherlands ranked at similar values of the stringency index [21], which seems to have led to a similar contact reduction.

Compared to the baseline, the effective number of contacts per person was found to be reduced by 69% in the April 2020 survey and by 0% in the June 2020 survey. This number would be proportional to the reproduction number (i.e. the number of secondary infections caused by a single infectious person in the population) under three conditions. First, the definition of contact (having a face-to-face conversation or physical contact) is a good proxy measure for at-risk contact events where SARS-CoV-2 can be transmitted. This condition is likely met as the virus transmits through similar routes as influenza virus, i.e. droplets, fomites, aerosols and contaminated surfaces, for which this is a validated approach [22]. Second, contacts should have been made in a similar fashion in the different surveys. Conversational contacts during the pandemic may very well have taken place at a larger distance or with a face mask. This would mean that the reproduction number would have been reduced further than by the reduction in the effective number of contacts. Third, all age groups are equally susceptible and infectious. As evidence accrues that children are less infectious or less susceptible [7, 23 – 25], and they have the largest numbers of contacts in the baseline survey, the reduction of the reproduction number is consequently less than the reduction in the effective number of contacts of 69% in the April 2020 survey. After relaxation of the measures, the focus of contact mixing shifts even more to younger age classes. Because of their limited role in transmission, the reduction of the effective reproduction number of COVID-19 should exceed the reduction in the effective number of contacts of 0%. Also general hygiene measures and use of face masks will have led to a reduction of the reproduction number, but this effect is not captured in the contact matrices.

The results of this study can immediately be applied to the public health response and management of the COVID-19 pandemic. The estimated contact reduction is applicable to other countries and regions with similar control measures. Combined with appropriate susceptibility and infectiousness profiles, the estimated age-specific contact matrices are useful for conducting scenario analyses with age-structured transmission models of COVID- 19, to project the future course of the epidemic with or without physical distancing measures [26 - 28]. As we plan to repeat the contact survey at regular intervals, we will be able to monitor the number of contacts made over time and the fraction of close contacts. With infection numbers on the rise again, we can detect the impact of renewed physical distancing measures, as well as changes in compliance. We believe that contact surveys such as these can help to inform and guide infection control measures in these times of unprecedented physical distancing.

## Supporting information

S2_contact_matrices

## Data Availability

Data of contact matrices shown in Fig. 4 are provided in tab separated file S2_contact_matrices.tsv.

## Acknowledgments

We gratefully acknowledge the participants of the contact surveys in 2016/2017 and 2020, as well as Dr. S. A. McDonald for critically assessing the manuscript.

## Supplementary material

Supplementary figures, tables and analyses are available in text file S1_supplement.pdf. Data of contact matrices shown in Fig. 4 are provided in tab separated file S2_contact_matrices.tsv.

