## Supplementary material for "The impact of physical distancing measures against COVID-19 transmission on contacts and mixing patterns in the Netherlands: repeated cross-sectional surveys in 2016/2017, April 2020 and June 2020": S1_supplement

"This supplementary material is hosted by *Eurosurveillance* as supporting information alongside the article The impact of physical distancing measures against COVID-19 transmission on contacts and mixing patterns in the Netherlands: repeated cross-sectional surveys in 2016/2017, April 2020 and June 2020, on behalf of the authors, who remain responsible for the accuracy and appropriateness of the content. The same standards for ethics, copyright, attributions and permissions as for the article apply. Supplements are not edited by *Eurosurveillance* and the journal is not responsible for the maintenance of any links or email addresses provided therein."

**S1.1 Household size distribution over age classes**

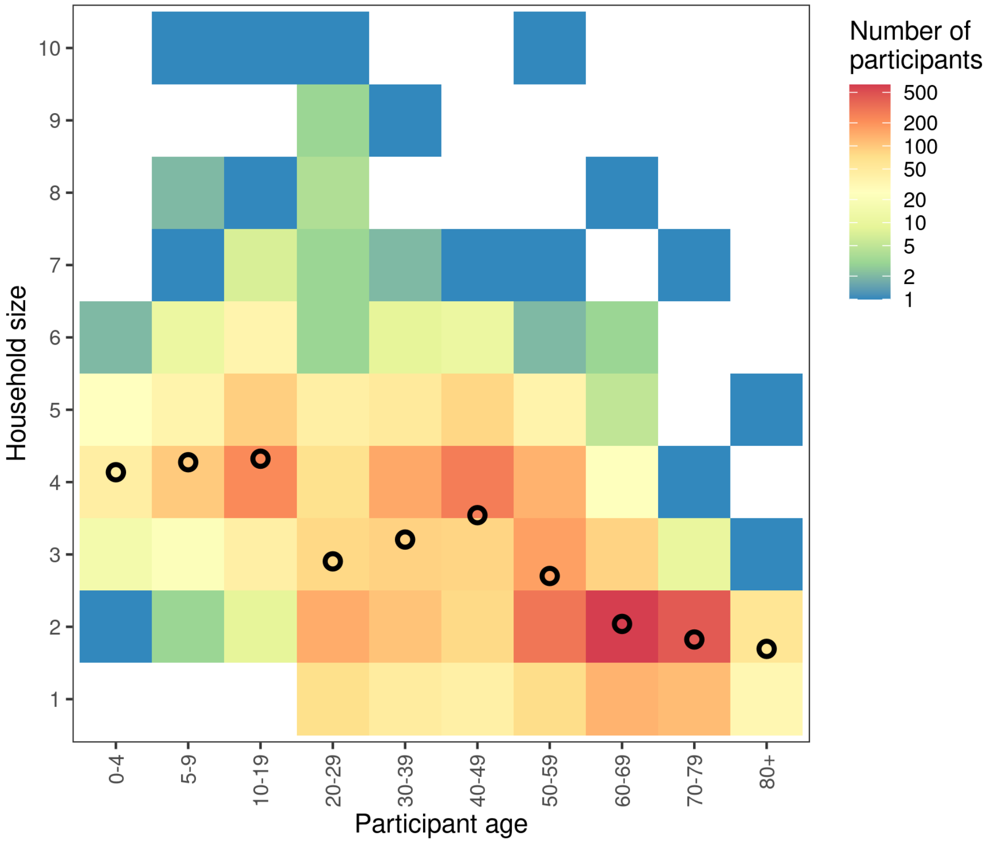


Figure S1 Household sizes reported by participants in het June 2020 survey, with frequency (color shadings) and the mean household size (black circle).

#### S1.2 Fraction of close community contacts

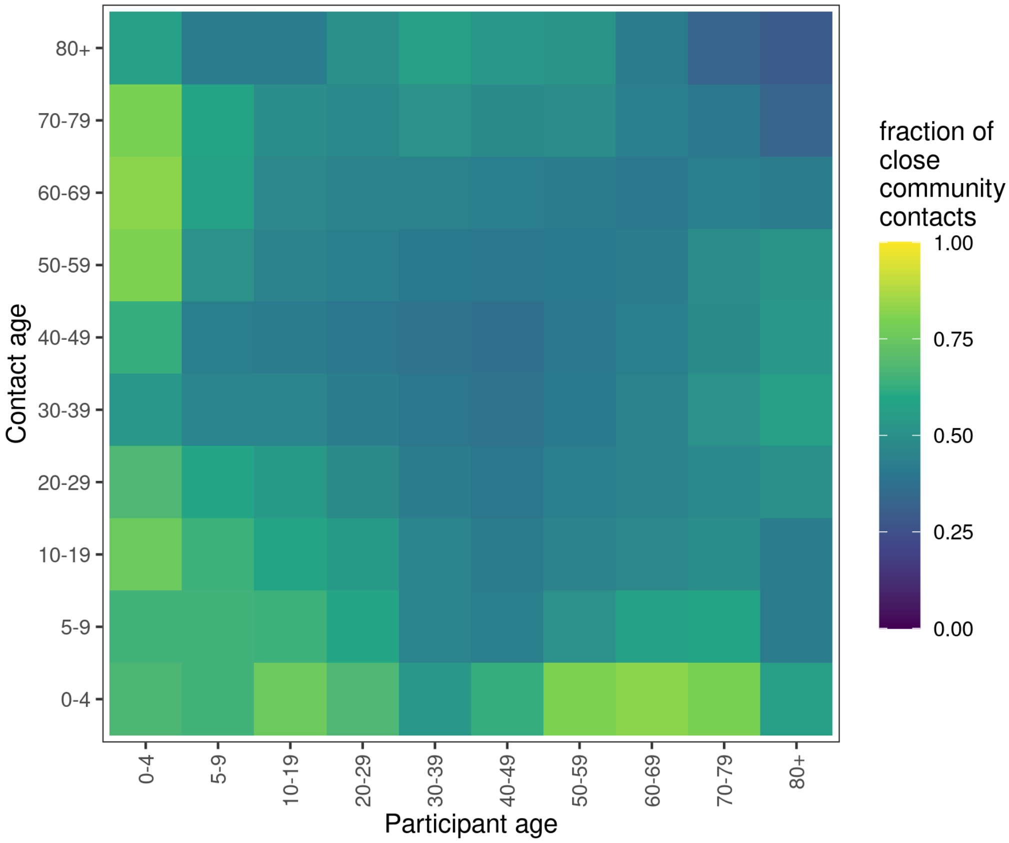


Figure S2 Fraction of close community contacts per participant between different age groups for the June 2020 survey. Close contacts were defined within 1.5 m.

**S1.3 Weighted number of community contacts per participant**

Table S1 Number of community contacts per participant in the baseline survey in 2016/2017, the April 2020 survey, and the June 2020 survey in the Netherlands (mean and interquartile range). Unweighted values (default) compared to values weighted values by participant age, participant sex, household size and contact day.

| | baseline | | April 2020 | | June 2020 | |
|---|---|---|---|---|---|---|
| | **mean** | **(IQR)** | **mean** | **(IQR)** | **mean** | **(IQR)** |
| Unweighted | 14.9 | (4 - 20) | 3.5 | (0 - 4) | 8.8 | (1 - 10) |
| Weighted by | | | | | | |
| Participant age | 14.5 | (4 - 19) | 3.4 | (0 - 4) | 9.7 | (1 - 13) |
| Participant sex | 14.9 | (4 - 20) | 3.5 | (0 - 4) | 8.8 | (1 - 10) |
| Household size | 14.9 | (4 - 20) | 3.5 | (0 - 4) | 8.8 | (1 - 11) |
| Contact day | 15.0 | (4 - 20) | 3.6 | (0 - 4) | 9.4 | (1 - 11) |

### S1.4 Number of community contacts per participant throughout the year

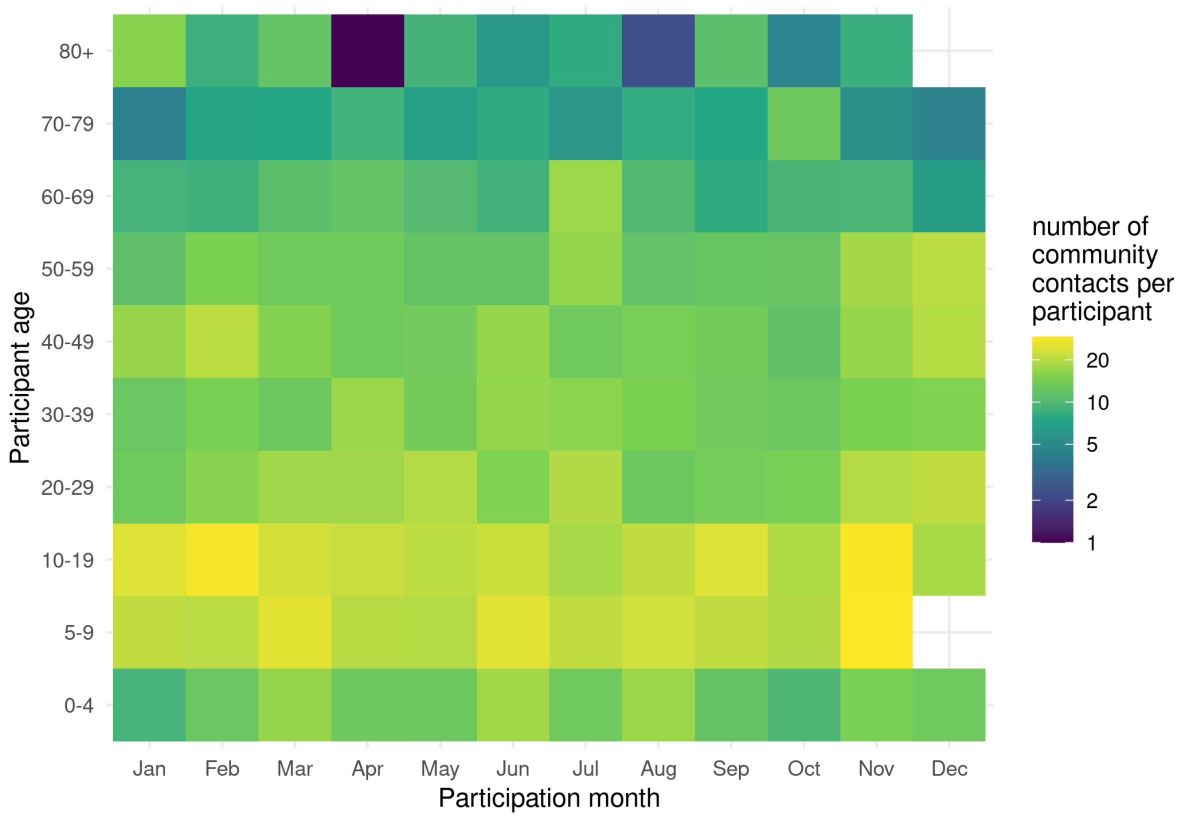


Figure S3 Number of community contacts per participant by age group throughout the year, as measured in baseline survey (2016/2017).

#### S1.5 Contact analysis of participants common to all surveys

In total 1739 participants were common to all three surveys. Their mean age was 40 years (range 0 – 86) in the baseline survey and 44 years (range 3 – 90) in the 2020 surveys. Also other characteristics of this group correspond to the participant characteristics of the full surveys (compare Fig S3 and Tab S2 to Fig 1 and Tab 1 in main text).

The percentage of these 1739 participants who did not report any community contacts increased from 4% (cf. 5%) in the baseline survey to 43% (cf. 42%) in the April 2020 survey, and decreased again to 21% (cf. 22%) in the June 2020 survey. The average number of community contacts a participant reported per day, decreased from 15.4 (cf. 14.9) in the baseline survey to 3.4 (cf. 3.5) in the April 2020 survey, and increased to 9.5 (cf. 8.8) in the June 2020 survey (Tab. S3). Also other contact reductions for the 1739 participants are similar to reductions found in the full surveys (compare Tab S3 and Fig S4 to Tab 2 and Fig 2 in main text).

The effective number of contacts per person (i.e. the largest eigenvalue of the contact matrix) is somewhat higher for the 1739 participants compared to the full survey, but shows the same trend: 21 (16 – 26, 95% credible interval) in the baseline survey, 5.8 (4.9 – 6.2) in the April 2020 survey, and 22 (14 – 31) in the June 2020 survey.

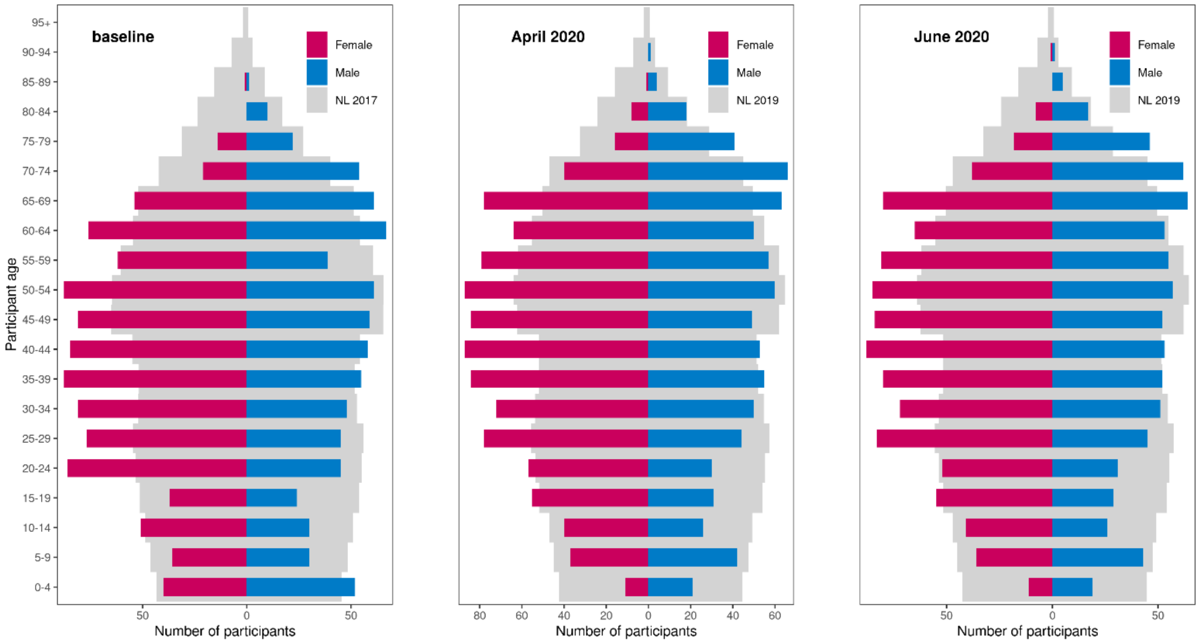


Figure S4 Composition according to age and sex of 1739 participants common to the baseline survey in 2016/2017, the April 2020 survey, and the June 2020 survey in the Netherlands; compared to the Dutch population in 2017, 2019 and 2019, respectively.

Table S2 Characteristics of participants common to the baseline survey in 2016/2017, the April 2020 survey, and the June 2020 survey in the Netherlands.

| | **baseline** | | **April 2020** | | **June 2020** | |
|---|---|---|---|---|---|---|
| | n | (%) | n | (%) | n | (%) |
| Total | 1739 | (100) | 1739 | (100) | 1739 | (100) |
| Participant age group | | | | | | |
| 0-4 | 92 | (5.3) | 32 | (1.8) | 30 | (1.7) |
| 5-9 | 66 | (3.8) | 79 | (4.5) | 79 | (4.5) |
| 10-19 | 142 | (8.2) | 152 | (8.7) | 151 | (8.7) |
| 20-29 | 253 | (14.5) | 209 | (12) | 211 | (12.1) |
| 30-39 | 272 | (15.6) | 261 | (15) | 255 | (14.7) |
| 40-49 | 283 | (16.3) | 273 | (15.7) | 277 | (15.9) |
| 50-59 | 250 | (14.4) | 283 | (16.3) | 278 | (16) |
| 60-69 | 258 | (14.8) | 255 | (14.7) | 262 | (15.1) |
| 70-79 | 111 | (6.4) | 163 | (9.4) | 164 | (9.4) |
| 80+ | 12 | (0.7) | 32 | (1.8) | 32 | (1.8) |
| Participant sex | | | | | | |
| Female | 978 | (56.2) | 978 | (56.2) | 978 | (56.2) |
| Male | 761 | (43.8) | 761 | (43.8) | 761 | (43.8) |
| Household size | | | | | | |
| 1 | 237 | (13.6) | 140 | (8.1) | 132 | (7.6) |
| 2 | 693 | (39.9) | 649 | (37.3) | 665 | (38.2) |
| 3 | 230 | (13.2) | 303 | (17.4) | 283 | (16.3) |
| 4 | 394 | (22.7) | 449 | (25.8) | 454 | (26.1) |
| 5 | 149 | (8.6) | 155 | (8.9) | 160 | (9.2) |
| 6+ | 36 | (2.1) | 43 | (2.5) | 45 | (2.6) |
| Contact day | | | | | | |
| Monday | 315 | (18.1) | 439 | (25.2) | 337 | (19.4) |
| Tuesday | 285 | (16.4) | 465 | (26.7) | 354 | (20.4) |
| Wednesday | 158 | (9.1) | 312 | (17.9) | 293 | (16.8) |
| Thursday | 80 | (4.6) | 170 | (9.8) | 172 | (9.9) |
| Friday | 151 | (8.7) | 83 | (4.8) | 88 | (5.1) |
| Saturday | 264 | (15.2) | 65 | (3.7) | 138 | (7.9) |
| Sunday | 344 | (19.8) | 203 | (11.7) | 356 | (20.5) |
| (Missing) | 142 | (8.2) | 2 | (0.1) | 1 | (0.1) |

Table S3 Number of community contacts per participant for 1739 participants common to the baseline survey in 2016/2017, the April 2020 survey, and the June 2020 survey in the Netherlands (mean and interquartile range).

| | **baseline** | | **April 2020** | | **June 2020** | |
|---|---|---|---|---|---|---|
| | **mean** | **(IQR)** | **mean** | **(IQR)** | **mean** | **(IQR)** |
| Total | 15.4 | (5 - 21) | 3.4 | (0 - 4) | 9.5 | (1 - 11) |
| Participant age | | | | | | |
| 0-4 | 12.2 | (3 - 19) | 1.9 | (0 - 3) | 22.8 | (6 - 32) |
| 5-9 | 27.0 | (9 - 36) | 2.1 | (0 - 3) | 28.5 | (8 - 40) |
| 10-19 | 23.4 | (9 - 34) | 3.1 | (0 - 4) | 16.5 | (2 - 26) |
| 20-29 | 18.3 | (6 - 25) | 3.3 | (0 - 5) | 9.8 | (1 - 12) |
| 30-39 | 15.3 | (5 - 19) | 4.1 | (0 - 5) | 10.3 | (2 - 13) |
| 40-49 | 16.8 | (6 - 22) | 4.5 | (0 - 4) | 9.5 | (2 - 12) |
| 50-59 | 13.1 | (4 - 17) | 4.6 | (0 - 6) | 6.3 | (1 - 8) |
| 60-69 | 9.9 | (3 - 12) | 2.2 | (0 - 3) | 5.9 | (0 - 6) |
| 70-79 | 8.7 | (2 - 12) | 1.8 | (0 - 2) | 2.5 | (0 - 4) |
| 80+ | 11.5 | (3 - 8) | 0.7 | (0 - 1) | 3.3 | (0 - 4) |
| Participant sex | | | | | | |
| Female | 15.4 | (5 - 20) | 3.1 | (0 - 4) | 9.5 | (1 - 11) |
| Male | 15.3 | (4 - 21) | 3.8 | (0 - 4) | 9.6 | (1 - 11) |
| Household size | | | | | | |
| 1 | 13.9 | (3 - 19) | 3.0 | (0 - 3) | 5.5 | (0 - 5) |
| 2 | 13.5 | (4 - 17) | 2.9 | (0 - 3) | 6.3 | (0 - 7) |
| 3 | 15.0 | (4 - 20) | 3.9 | (0 - 5) | 10.1 | (2 - 13) |
| 4 | 17.6 | (6 - 23) | 3.7 | (0 - 4) | 12.0 | (2 - 15) |
| 5 | 19.1 | (7 - 28) | 3.9 | (0 - 4) | 15.3 | (1 - 21) |
| 6+ | 23.9 | (10 - 34) | 3.8 | (0 - 5) | 20.7 | (2 - 29) |
| Contact day | | | | | | |
| Monday | 18.0 | (5 - 25) | 3.1 | (0 - 4) | 11.3 | (2 - 12) |
| Tuesday | 16.5 | (6 - 23) | 3.4 | (0 - 4) | 9.1 | (1 - 11) |
| Wednesday | 16.7 | (5 - 20) | 3.7 | (0 - 4) | 11.4 | (2 - 14) |
| Thursday | 19.4 | (6 - 34) | 4.4 | (0 - 5) | 11.3 | (2 - 15) |
| Friday | 15.4 | (5 - 20) | 4.0 | (0 - 5) | 12.2 | (2 - 13) |
| Saturday | 13.2 | (4 - 17) | 4.1 | (0 - 4) | 8.3 | (2 - 9) |
| Sunday | 11.5 | (3 - 14) | 2.4 | (0 - 3) | 5.7 | (0 - 5) |
| (Missing) | 17.1 | (6 - 24) | 3.0 | (3 - 3) | 2.0 | (2 - 2) |

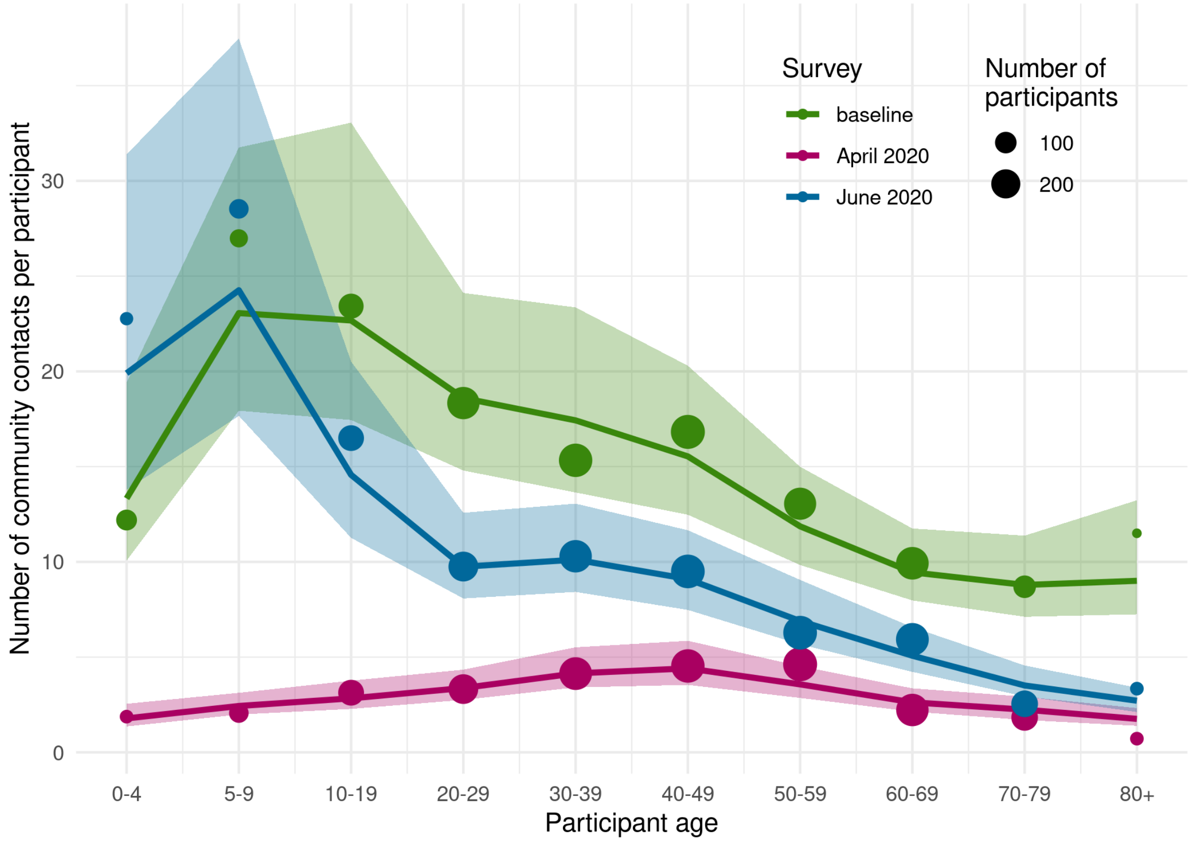


Figure S5 Number of community contacts per participant per age group for 1739 participants common to the baseline survey in 2016/2017 (green), the April 2020 survey (red), and the June 2020 survey (blue) in the Netherlands; shown are the estimated mean numbers of contacts after accounting for reciprocity in contacts (line) with the corresponding 95% credible interval (shaded area), and the observed means (size of points scaled to number of participants).
